# Prediction of peak oxygen consumption using cardiorespiratory parameters from warm-up and submaximal stage of treadmill cardiopulmonary exercise test

**DOI:** 10.1101/2023.09.06.23295118

**Authors:** Maciej Rosoł, Monika Petelczyc, Jakub S. Gąsior, Marcel Młyńczak

## Abstract

This study investigates the quality of peak oxygen consumption (VO_2peak_) prediction based on cardiac and respiratory parameters calculated from warmup and submaximal stages of treadmill cardiopulmonary exercise test (CPET) using machine learning (ML) techniques and assesses the importance of respiratory parameters for the prediction outcome. The database consists of the following parameters: heart rate (HR), respiratory rate (RespRate), pulmonary ventilation (VE), oxygen consumption (VO_2_) and carbon dioxide production (VCO_2_) obtained from 369 treadmill CPETs. Combinations of features calculated based on the HR, VE and RespRate time-series from different stages of CPET were used to create 11 datasets for VO_2peak_ prediction. Thirteen ML algorithms were employed, and model performances were evaluated using cross-validation with mean absolute percentage error (MAPE), R^2^ score, mean absolute error (MAE), and root mean squared error (RMSE) calculated after each iteration of the validation. The results demonstrated that incorporating respiratory-based features improves the prediction of VO_2peak_. The best results in terms of R^2^ score (0.47) and RMSE (5.78) were obtained for the dataset which included both cardiac- and respiratory-based features from CPET up to 85% of age-predicted HR_max_, while the best results in terms of MAPE (10.5%) and MAE (4.63) were obtained for the dataset containing cardiorespiratory features from the last 30 seconds of warmup. The study showed the potential of using ML models based on cardiorespiratory features from submaximal tests for prediction of VO_2peak_ and highlights the importance of the monitoring of respiratory signals, enabling to include respiratory parameters into the analysis. Presented approach offers a feasible alternative to direct VO_2peak_ measurement, especially when specialized equipment is limited or unavailable.

## 1. Introduction

Peak oxygen consumption (VO_2peak_) obtained through cardiopulmonary exercise test (CPET) is the gold standard measure of cardiorespiratory fitness (1). It is a reliable predictor of cardiac events, as well as lung cancer and liver transplantation survival and risk of postoperative complications (2–6). Moreover, VO_2peak_ is a predictor of sport performance (7–9) and planetary mission task performance during spaceflight (10). Although CPET is the most reliable form of test, it is costly, requires specialized personnel and advanced equipment (11).

While conducting CPET, heart rate (HR) data are usually obtained through electrocardiography (ECG), while respiratory rate (RespRate) and pulmonary ventilation (VE) are gathered using tight-fitting masks. Nevertheless, this data can be acquired with relative ease, using heart rate monitors or smartwatches in case of HR, and impedance pneumography (IP) in case of RespRate and VE (12,13). Moreover, CPET is physically demanding as assumes the participants’ exhaustion and thus it is contraindicated for patients with acute myocardial infarction, unstable angina, uncontrolled arrhythmia causing symptoms or hemodynamic compromise, uncontrolled asthma, and other pathological conditions (11). Maximal cardiopulmonary exercise test might also interfere with an athletes training program (14).

Actually, thanks to: a) the growing development of aforementioned measurement devices, b) availability of simply field-based tests such as incremental shuttle walk test (15,16) and c) new statistical prediction models and equations, clinicians and/or researchers are able to estimate VO_2peak_, and/or VO_2max_, based on selected parameters without performing maximal CPET (17–22). Unfortunately, estimated VO_2peak_ using, e.g., only 6-min Walk Test distance demonstrated poor agreement with measured VO_2peak_ from a CPET (23). Addition of other data such as demographic, anthropometric, and functional characteristics improved the accuracy of VO_2peak_ estimate based on walking tests at least in elderly patients with stable coronary artery disease (model with all variables explained 73% of VO_2peak_ variance) (24). Thus, estimation of peak oxygen consumption based on combination of demographic factors and cardiac parameters obtained during submaximal (or even not) physical effort is possible, however, may be biased.

Reliable and accurate estimation of VO_2peak_ without performing maximal CPET may require more input physiological data to perform more sophisticated analyses. Thus, the development of new prediction models or equations, which will be able to accurately estimate VO_2peak_, and/or VO_2max_, and will not relies on performing maximal CPET, is still ongoing (18,25). In recent years with the growth of the popularity of machine learning tools (ML) incorporated during the data analysis phase, those techniques were also utilized for the prediction of VO_2_ kinetics and VO_2max_ (26,27). ML models were also used by *Szijarto et al*. for prediction of VO_2peak_ based on the anthropometric data and 2D echocardiography (2DE) (28). This approach was more accurate than a model based on anthropometric factors, however, it required performing a 2DE examination with sophisticated equipment and a trained physician. Importantly, not only the model or prediction algorithm might be important in terms of the prediction accuracy, but also the features used for the training. There are existing studies utilizing respiratory rate and ML for prediction of oxygen uptake dynamics during CPET (29–31). However, to the best of our knowledge, there have been no previous studies utilizing features from cardiorespiratory time-series obtained from submaximal CPET, for the prediction of VO_2peak_ using ML models.

The aim of this paper was hence to investigate the quality of VO_2peak_ prediction by models based on cardiac and respiratory features obtained from different stages of CPET. Additionally, we assessed the importance of respiratory-based features included in the models for VO_2peak_ prediction.

## 2. Materials and Methods

### 2.1. Data and study population

The database of cardiorespiratory time-series acquired during treadmill maximal cardiopulmonary exercise tests presented by *Mongin et al*. was used (32,33). The database comprises 992 recordings from experiments undertaken among amateur and professional athletes in the Exercise Physiology and Human Performance Lab of the University of Málaga between 2008 and 2018 with two types of protocols: a continuous increase of treadmill speed and a graded approach. The test itself was preceded by a warmup at 5 km/h. The study was conducted according to the principles of the Declaration of Helsinki, the study protocol was approved by the Research Ethics Committee of the University of Málaga, written informed consent was obtained from the participants and all the data were analyzed anonymously.

During each test, the following cardiorespiratory time-series were acquired: heart rate (HR), respiratory rate (RespRate), pulmonary ventilation (VE), oxygen uptake (VO_2_) and carbon dioxide production (VCO_2_). Data were acquired on a breath-to-breath basis. HR was monitored via a 12-lead ECG (Mortara Instrument, Inc., USA), while respiratory signals were obtained using the CPX MedGraphics gas analyzer system (Medical Graphics Corporation, USA) (32).

Participants between 18 and 40 years old were chosen for the analysis reducing the sample size to 692. Tests only with ramp speed increments were selected in order to obtain more consistent conditions along the study population. In result, 485 recordings have left. Next, subjects who were determined as outliers based on the 1.5 interquartile range method in terms of weight, height, and VO_2peak_, with respect to the given sex, were excluded from the study, limiting to 462 recordings. Furthermore, the obtained data was visually evaluated in order to discard measurements during which there were visible artefacts in HR acquisition (e.g., sudden drop of over 30 bpm or lack of continuity of HR time-series during CPET probably due to electrode detachment); ultimately 369 recordings became background for the analysis. The final recordings belong to 327 unique subjects (42 subjects had more than one test) including 275 men and 52 women. The demographic summary of the final group is presented in Table 1.

**Table 1.**
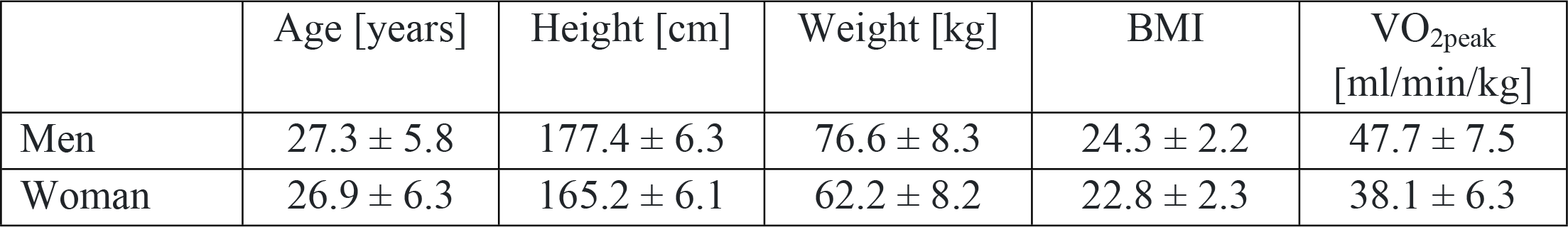
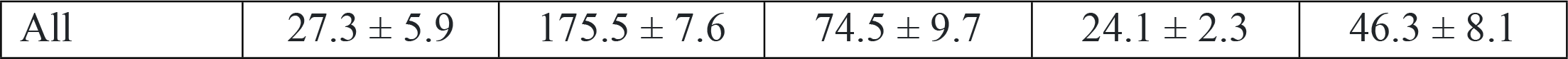
Descriptive statistics of the study population.

| | Age [years] | Height [cm] | Weight [kg] | BMI | $\text{VO}_{2\text{peak}}$<br>[ml/min/kg] |
| --- | --- | --- | --- | --- | --- |
| Men | 27.3 ± 5.8 | 177.4 ± 6.3 | 76.6 ± 8.3 | 24.3 ± 2.2 | 47.7 ± 7.5 |
| Woman | 26.9 ± 6.3 | 165.2 ± 6.1 | 62.2 ± 8.2 | 22.8 ± 2.3 | 38.1 ± 6.3 |
| All | $27.3 \pm 5.9$ | $175.5 \pm 7.6$ | $74.5 \pm 9.7$ | $24.1 \pm 2.3$ | $46.3 \pm 8.1$ |

### 2.2. Modeling

Based on the aforementioned dataset, we decided to investigate the quality of VO_2peak_ prediction from different stages of CPET based on cardiac and respiratory parameters, and to assess the importance of respiratory-based features included in the modeling of VO_2peak_. For this purpose, we utilized recorded time-series of HR, RespRate, and VE. VO_2peak_ was determined as the maximal value of the signal obtained after a 15-breath VO_2_ moving average window according to the recommendation presented by *Robergs et al*. (34).

As features for ML models, basic statistics such as mean, standard deviation, maximal and minimal value, median, 25^th^ and 75^th^ quantile, skewness, kurtosis, coefficient from linear regression, impulse and shape factors were calculated for HR, RespRate, and VE, for a given stage of the maximal CPET. On this basis, 11 datasets were created based on different combinations of parameters and CPET stages, as presented in Table 2. Our research is focused on the submaximal stage from the cardiopulmonary exercise test, which equals 85% of the maximal measured and age-predicted HR_max_ as a threshold. Studied value of HR termination is commonly used in submaximal testing (35–37). We also used both actual HR_max_ obtained during the treadmill cardiopulmonary exercise test, and age-predicted HR_max_ (220-age) in order to provide insights about the utility of the prediction of VO_2peak_ in submaximal tests without prior knowledge about the value of HR_max_ for a given subject. The example plot of the signals, alongside the threshold for all the stages of the CPET for which the features were calculated, is presented in Figure 2.

**Table 2.** Characteristics of all datasets with an indication of features belonging to individual datasets.

| Dataset | subjects' demography | HR features from the last 30 s of warmup | RespRate and VE features from the last 30 s of warmup | HR features from CPET up to 85% of $HR_{max}$ | RespRate and VE features from CPET up to 85% of $HR_{max}$ | HR features from CPET up to 85% of age-predicted $HR_{max}$ | RespRate and VE features from CPET up to 85% of age-predicted $HR_{max}$ |
| --- | --- | --- | --- | --- | --- | --- | --- |
| D1 | + |  |  |  |  |  |  |
| D2 | + | + |  |  |  |  |  |
| D3 | + | + | + |  |  |  |  |
| D4 | + |  |  | + |  |  |  |
| D5 | + |  |  | + | + |  |  |
| D6 | + | + |  | + |  |  |  |
| D7 | + | + | + | + | + |  |  |
| D8 | + |  |  |  |  | + |  |
| D9 | + |  |  |  |  | + | + |
| D10 | + | + |  |  |  | + |  |
| D11 | + | + | + |  |  | + | + |

**Figure 1.**
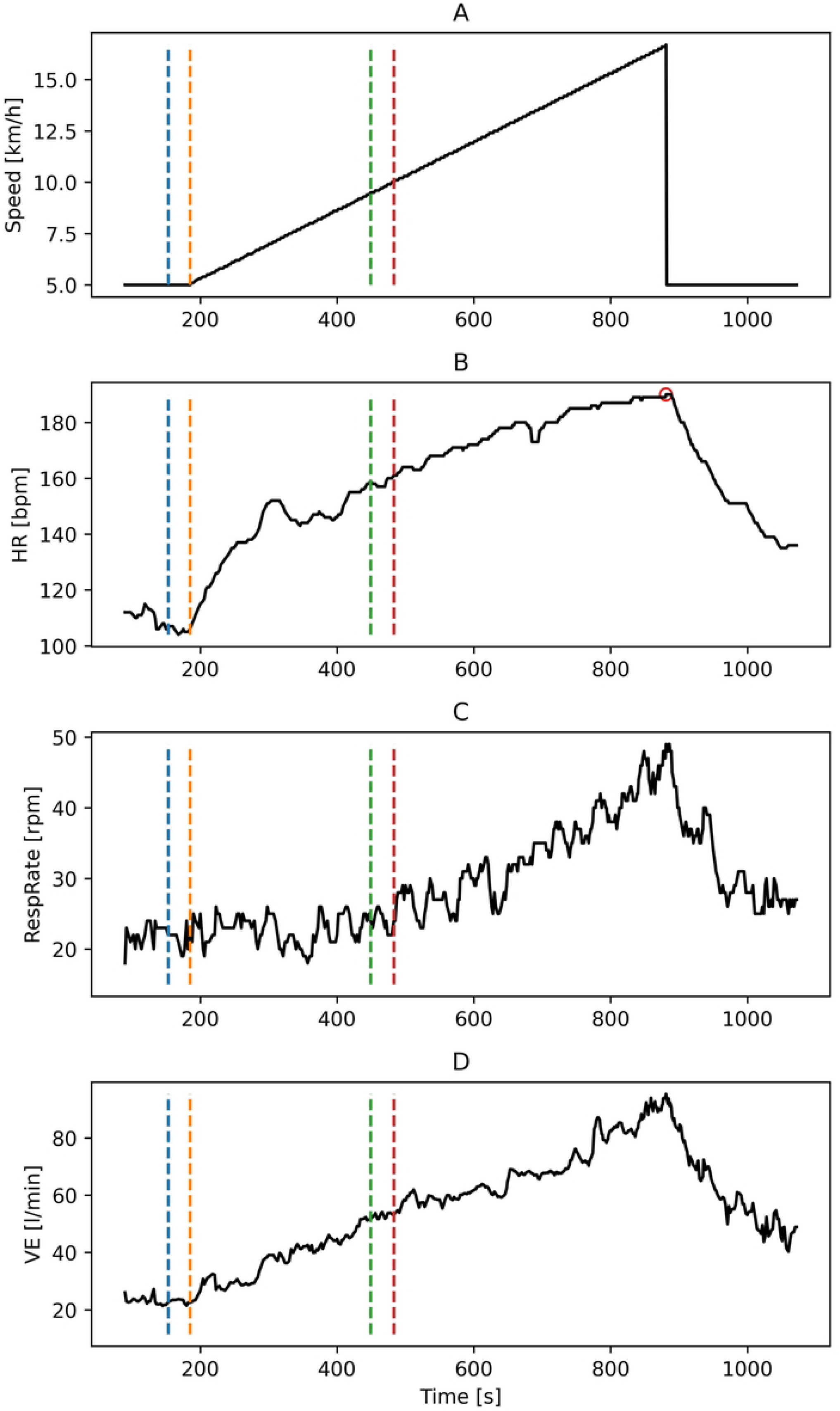
Typical representation of the time-series for participants with selected fragments used in the analysis. Part A presents the linearly increasing treadmill speed, part B heart rate fluctuations, part C respiratory rate and part D pulmonary ventilation kinetics. The segment between the blue and orange dashed lines is the last 30 seconds of warmup. The segment between the orange and green lines corresponds to the section of CPET up to 85% of the age-predicted HR_max_. Finally, the segment between the orange and red lines corresponds to the increasing workload in CPET up to 85% of the measured HR_max_, which is marked with a red circle on the HR plot.

**Figure 2.**
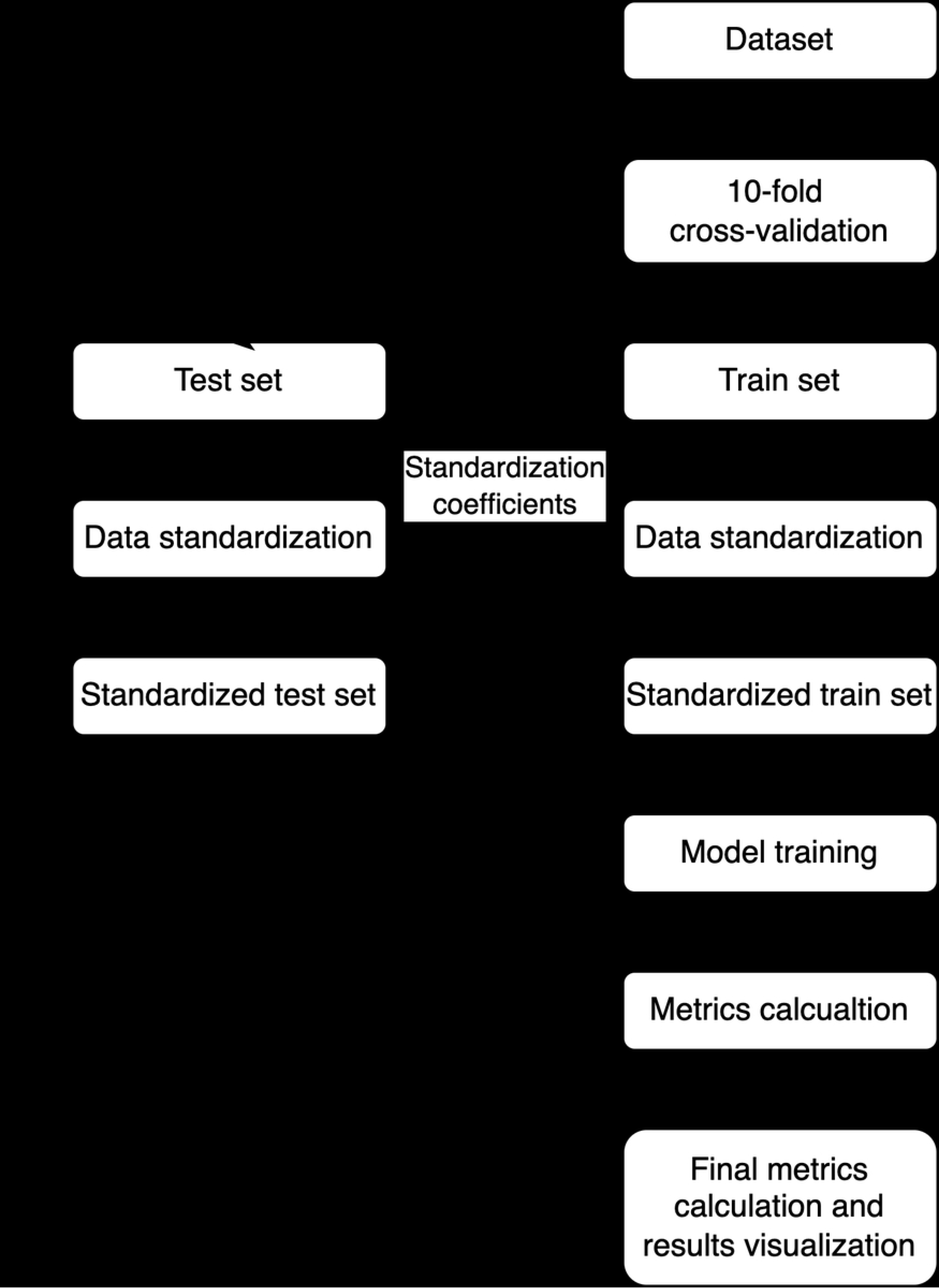
Modeling pipeline applied for each dataset and algorithm.

The 10-fold cross-validation (CV) was used to assess the accuracy of the prediction. In each iteration, standardization of the non-categorical features based on the mean and standard deviation from the training dataset was performed. The only feature that was not standardized was participants’ sex: -1 was assigned to male, and 1 to female subjects. Different ML algorithms were utilized: Linear, Lasso and Ridge Regression, Random Forest, XGBoost, Multilayer perceptron, Epsilon-Support Vector Regression, Bayesian Ridge Regression, Bayesian Automatic Relevance Determination (ARD) Regression, Gaussian Process Regression, Gradient Boosting for Regression, Huber Regression and Theil-Sen Estimator (38–40). The hyperparameter tuning was performed for each algorithm using the grid-search technique. In each iteration of the validation, metrics like mean absolute percentage error (MAPE), R^2^ score, mean absolute error (MAE) and root mean squared error (RMSE) were calculated. The best model for each dataset was determined based one the lowest MAPE score (which was chosen arbitrarily) obtained from the cross-validation. For the best model, Lin concordance correlation coefficient was calculated and results were visualized as the dependency between predicted and actual values of VO_2peak_ and as Bland-Altman plot.

Metrics obtained from all datasets were pairwise compared using the Wilcoxon signed-rank test. The significance level was set to 0.05. For the calculations, Python 3.9.13 was used. The whole modeling pipeline is presented in Figure 2.

### 2.3. Explainable AI

In order to investigate the importance of the individual features used for ML modeling, explainable artificial intelligence (XAI) tools were applied. For this purpose, the Dalex Python package was used (41). During each iteration of the cross-validation, Shapley values and model-level variable importance based on drop-out loss values were calculated on the test set. After the whole cross-validation, all Shapley values for each sample and feature, as well as mean variable importance values were visualized. For the variable importance, *model*_*parts* function of *dalex*.*Explainer* class was used. 30 permutation rounds were performed on each variable with MAE as a loss function and no data sampling (argument *N* was equal to *None*) due to the small number of samples.

## 3. Results

The metrics obtained for the best algorithm in terms of the lowest MAPE from the cross-validation for each dataset are presented in Table 3 alongside the model names. The violin-plots of the obtained metrics for each dataset were visualized in Figure 3. The p-values from the Wilcoxon signed-rank test from a pairwise comparison of the metrics are presented in Figure 3.

**Table 3.**
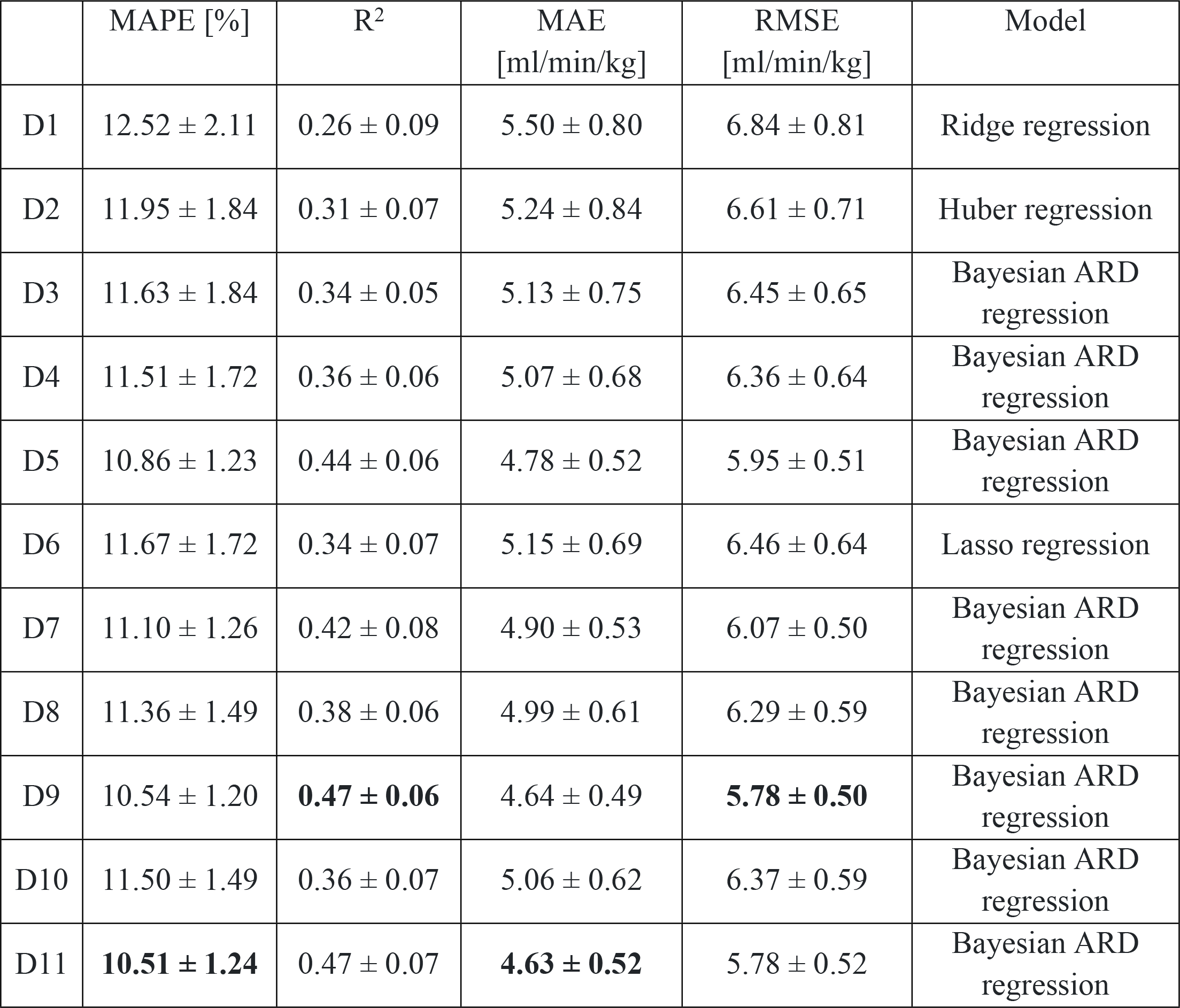
Mean and standard deviation of metrics from cross-validation for each dataset for the model which resulted in the lowest MAPE for the given dataset. The highest metric values were highlighted.

|  | MAPE [%] | R<sup>2</sup> | MAE<br>[ml/min/kg] | RMSE<br>[ml/min/kg] | Model |
| --- | --- | --- | --- | --- | --- |
| D1 | 12.52 ± 2.11 | 0.26 ± 0.09 | 5.50 ± 0.80 | 6.84 ± 0.81 | Ridge regression |
| D2 | 11.95 ± 1.84 | 0.31 ± 0.07 | 5.24 ± 0.84 | 6.61 ± 0.71 | Huber regression |
| D3 | 11.63 ± 1.84 | 0.34 ± 0.05 | 5.13 ± 0.75 | 6.45 ± 0.65 | Bayesian ARD<br>regression |
| D4 | 11.51 ± 1.72 | 0.36 ± 0.06 | 5.07 ± 0.68 | 6.36 ± 0.64 | Bayesian ARD<br>regression |
| D5 | 10.86 ± 1.23 | 0.44 ± 0.06 | 4.78 ± 0.52 | 5.95 ± 0.51 | Bayesian ARD<br>regression |
| D6 | 11.67 ± 1.72 | 0.34 ± 0.07 | 5.15 ± 0.69 | 6.46 ± 0.64 | Lasso regression |
| D7 | 11.10 ± 1.26 | 0.42 ± 0.08 | 4.90 ± 0.53 | 6.07 ± 0.50 | Bayesian ARD<br>regression |
| D8 | 11.36 ± 1.49 | 0.38 ± 0.06 | 4.99 ± 0.61 | 6.29 ± 0.59 | Bayesian ARD<br>regression |
| D9 | 10.54 ± 1.20 | **0.47 ± 0.06** | 4.64 ± 0.49 | **5.78 ± 0.50** | Bayesian ARD<br>regression |
| D10 | 11.50 ± 1.49 | 0.36 ± 0.07 | 5.06 ± 0.62 | 6.37 ± 0.59 | Bayesian ARD<br>regression |
| D11 | **10.51 ± 1.24** | 0.47 ± 0.07 | **4.63 ± 0.52** | 5.78 ± 0.52 | Bayesian ARD<br>regression |

**Figure 3.**
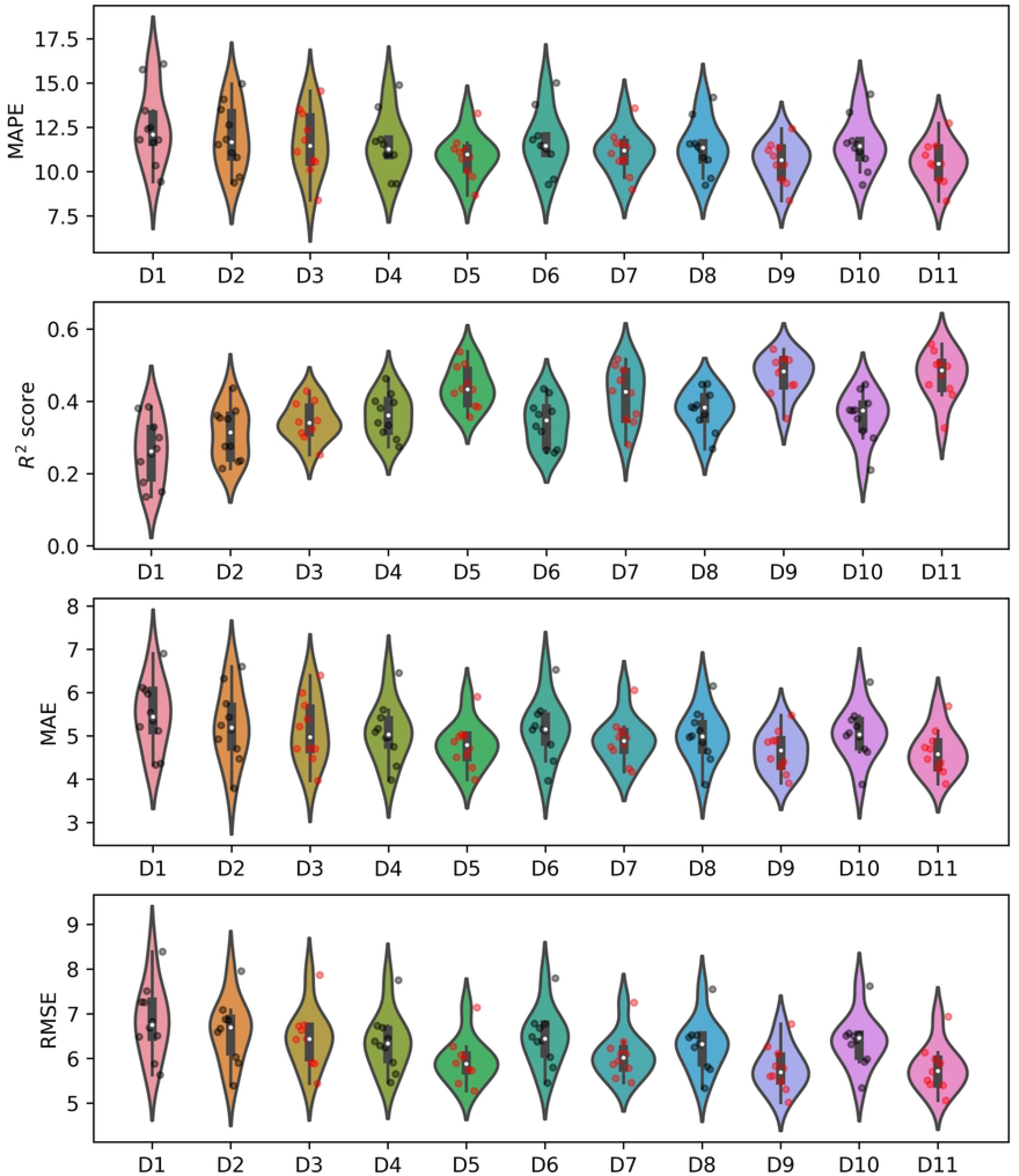
Violin-plots of the calculated metrics for each dataset with the visualization of the metrics obtained in each iteration of the 10-fold cross-validation. Black dots represent metrics obtained from datasets without respiratory-based features, while red dots represent these that include such features.

**Figure 4.**
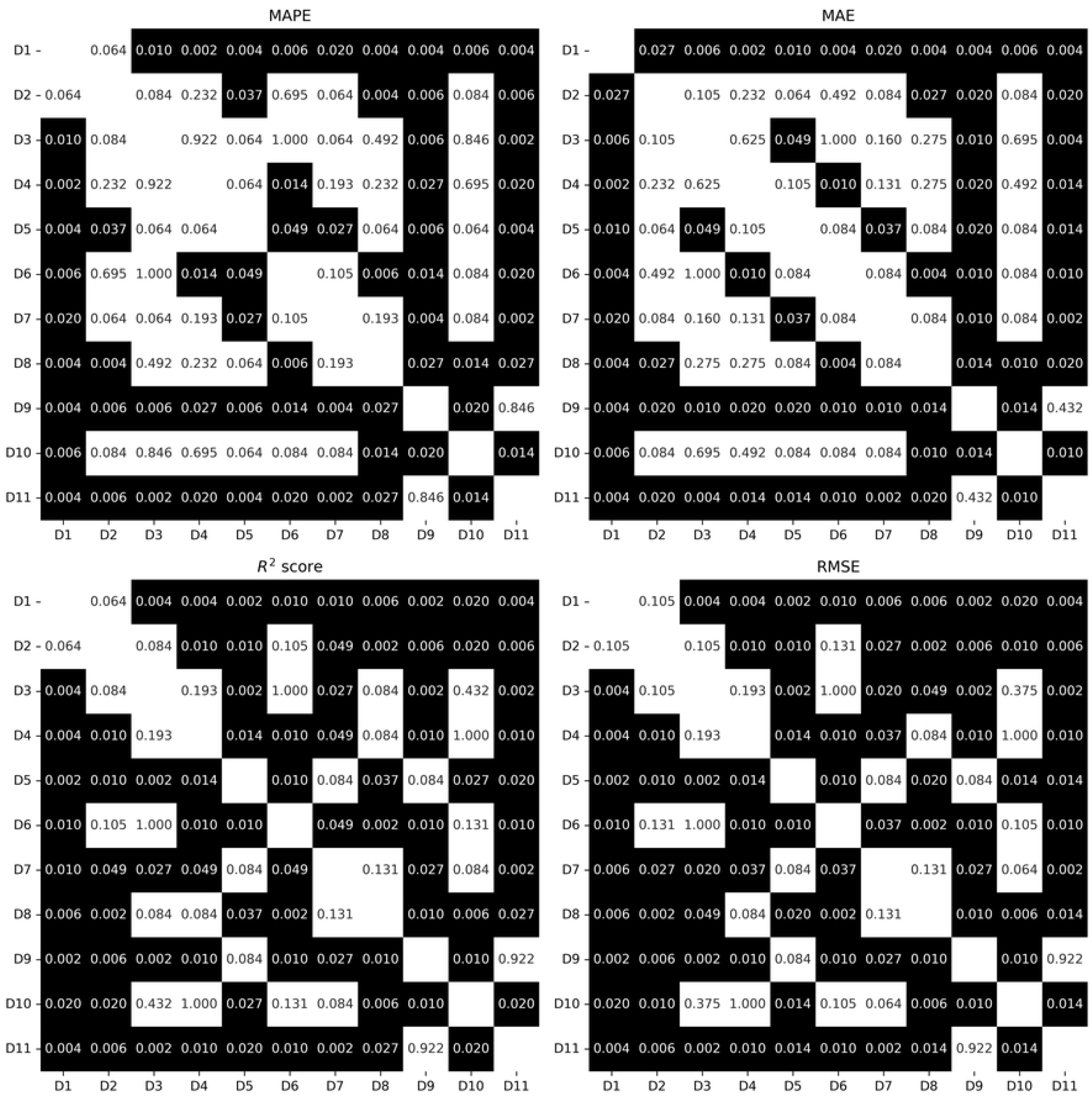
The p-values from Wilcoxon signed-rank test from pairwise comparison of the metrics obtained from different datasets. P-values smaller than 0.05 are marked with a black background.

The lowest MAPE and MAE - 10.51% and 4.63, respectively - were obtained for dataset D11 (demographic data along with cardiac and respiratory features from the last 30 seconds of warmup and CPET up to 85% of age-predicted HR_max_), while the lowest RMSE and highest R^2^ score (5.78 and 0.47, respectively) were obtained for D9 (demographic data along with cardiac and respiratory features from CPET up to 85% of age-predicted HR_max_). The worst prediction of VO_2peak_ in terms of all metrics was achieved by using the D1 (demographic data) dataset. Results obtained for D11 were statistically significantly better in terms of all metrics than results for all the rest of the datasets excluding D9 as presented in Figure 3. Regarding R^2^ score and RMSE metrics, datasets that included respiratory-based features from the part of CPET (irrespective of HR_max_ determination, whether measured or estimated) showed statistically significant superiority over datasets lacking features based on VE and respiratory rate during the corresponding period as presented in Figure 3. Similarly, for MAPE and MAE, datasets containing respiratory-based features calculated up to 85% of age-predicted HR_max_ demonstrated significantly better metrics than datasets without such features.

The measured values of VO_2peak_ and values predicted for the dataset that obtained the lowest MAPE score (D11) were visualized in Figure 5. The Lin concordance correlation coefficient between predicted and measured VO_2peak_ values was 0.66. The Bland-Altman plot for this dataset is presented in Figure 6.

**Figure 5.**
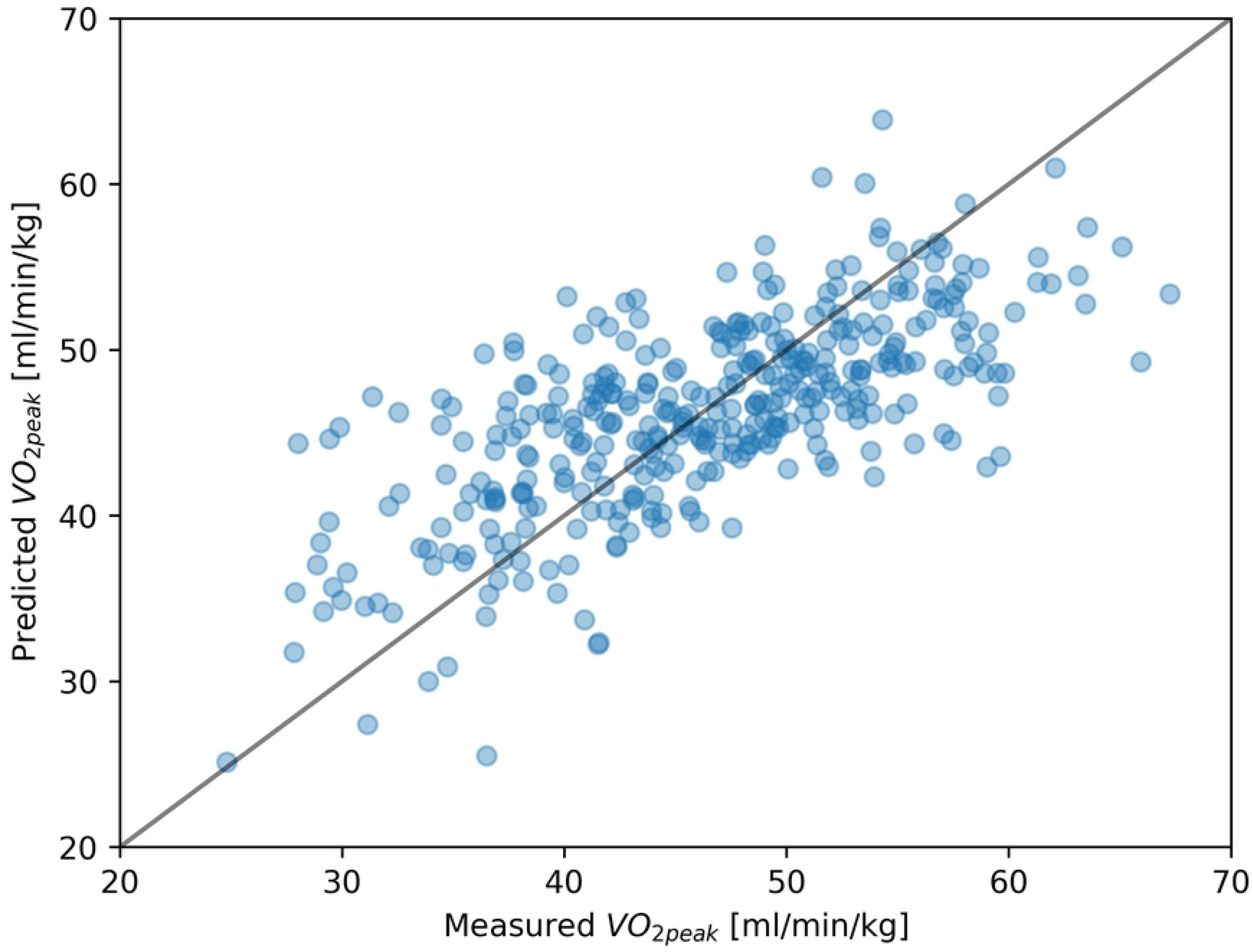
The plot of measured and predicted VO_2peak_ values for dataset D11. The solid black line represents the function where predicted value is equal to the measured one.

**Figure 6.**
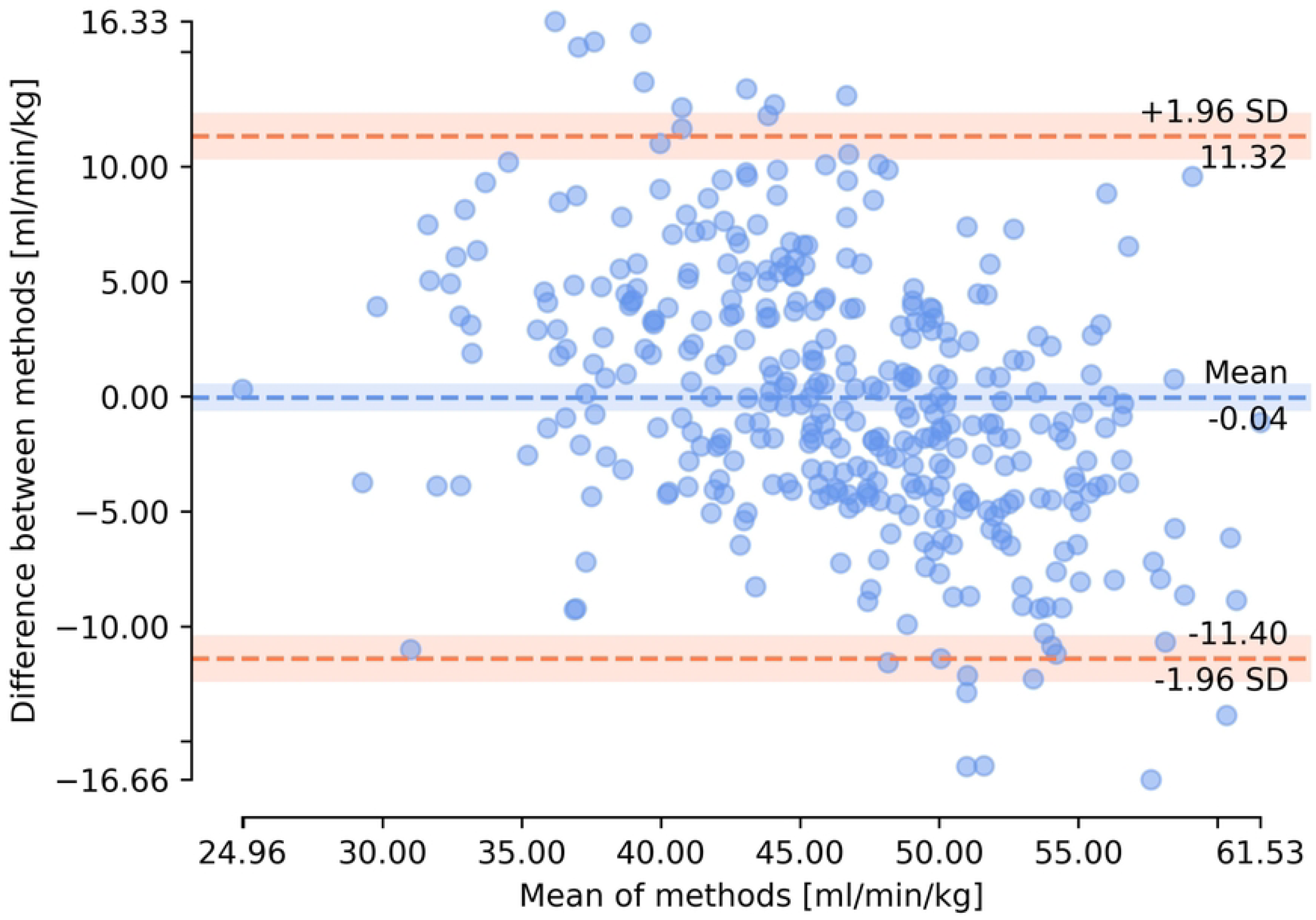
Bland-Altman plot of measured (gold standard from CPET) and predicted VO_2peak_ values based on the results for dataset D11.

As the smallest mean MAPE was obtained for D11, Shapley values and feature importance were visualized for this dataset in Figures 7 and 8, respectively. The discussion of the XAI results can be found in the next section.

**Figure 7.**
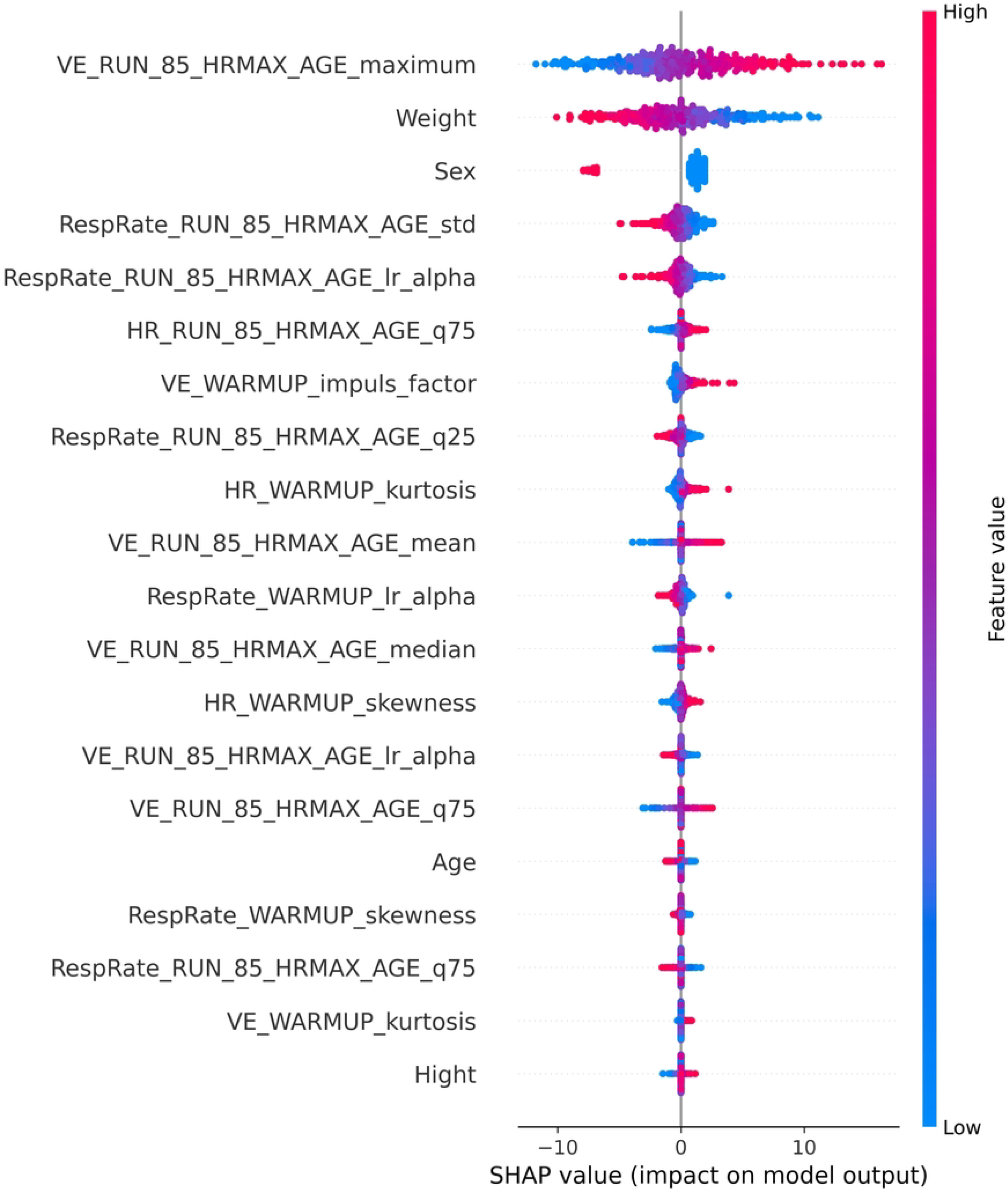
Shapley values obtained for dataset D11. Feature names are explained in the Appendix 1.

**Figure 8.**
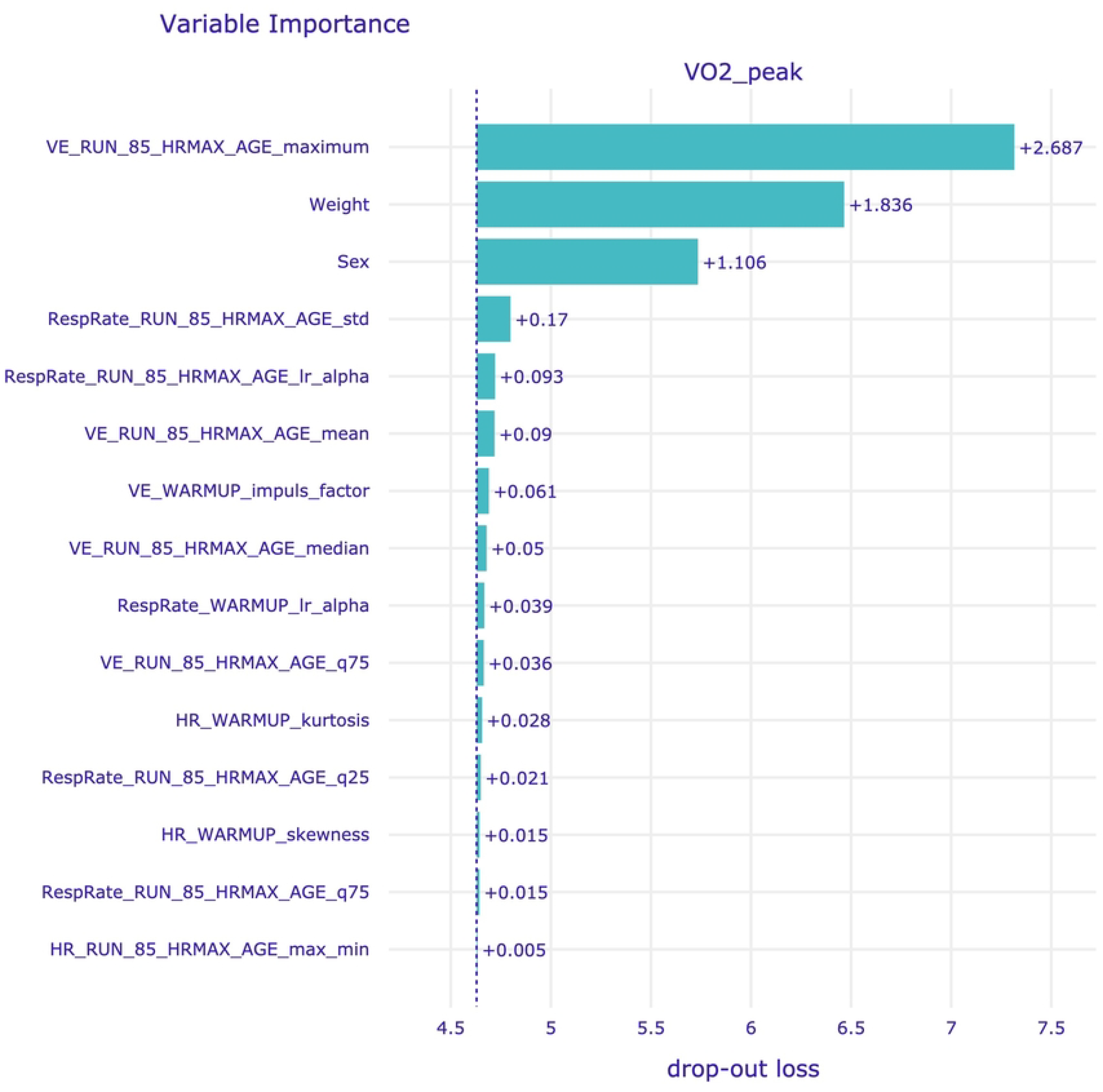
Variable importance for dataset D11. Feature names are explained in the Appendix 1.

## 4. Discussion

Considering the features calculated from HR, VE, and RespRate time-series (attainable without the specialized equipment used in CPET), it is possible to predict VO_2peak_ from a submaximal test relying on age-predicted HR_max_, achieving a mean absolute percentage error of 10.51% (for D11), using Bayesian ARD regression method. The addition of respiratory-based parameters resulted in an improvement of prediction compared to datasets based solely on the corresponding stage of the treadmill cardiopulmonary exercise test in 4 out of 5 cases in terms of R^2^ score and RMSE, and 2 out of 5 cases in terms of MAPE and MAE. When limiting treadmill cardiopulmonary exercise test to 85% of age-based HR_max_, the inclusion of features based on VE and RespRate improved the prediction in terms of all the specified metrics. The fact that the best results were achieved for the dataset considering 85% aged-based HR_max_ and parameters obtained from easily accessible time-series indicates the possibility of using the presented method in clinical practice to determine VO_2peak_ without the prior knowledge of the actual HR_max_ value and the necessity to perform a maximal treadmill cardiopulmonary exercise test.

Obtaining VO_2peak_ from maximal CPET might be costly, time-consuming and in some cases impossible or contraindicated to carried out due to observed cardiac or pulmonary dysfunction, musculoskeletal diseases, or strict training programs. Therefore, there is a growing interest in the prediction of VO_2peak_ and/or VO_2max_ from submaximal tests (14,42–48). Our study focused on investigating ML algorithms to predict VO_2peak_ with the set of features, which could be obtained using simpler techniques than commonly used spirometry, and the significance of incorporating respiration into the prediction process. The presented results are similar or superior compared to some other presented VO_2peak_ prediction methods like WFI VO_2peak_ prediction equation, deep-learning model based on 2DE, or regression models from PACER 20-m shuttle run (19,28,49–52). However, in the existing literature, there are also techniques, which managed to obtain better performance like regression models based on submaximal exercise test protocol using a total body recumbent stepper (53–55). Nonetheless, in those studies more heterogeneous groups of patients were present in terms of age or health status (patients after heart failure or individuals with low to moderate risk of cardiovascular diseases). Further improvement of the prediction of VO_2peak_ might be achieved by increasing sample size, and inclusion of other parameters based on the raw signals (especially ECG) like HRV and parameters from information and causal domain (56–59).

Another notable aspect of the study was the utilization of XAI tools, specifically Shapley values and model-level variable importance, to obtain insights into the feature importance for prediction. For most datasets (including D9 and D11, which produced the best results) Bayesian ARD Regression model was used, which has an ability to automatically determine the relevance of each feature, effectively pruning irrelevant or redundant information, while accentuating the impactful variables (60). In our analysis, we found that the top five most influential features were consistent between Shapley values and variable importance. The most impactful feature of the prediction was the maximal value of VE during the test, up to 85% of age-predicted HR_max_. Additionally, subjects’ weight and sex influenced the prediction results, with higher VO_2peak_ observed in lighter individuals and males compared to females. Notably, 13 out of the 20 features with the highest Shapley values and 10 out of the 15 features with the highest variable importance score were related to respiratory signals. Those findings seem to be in line with results presented in other studies, where the importance of respiratory signals in the context of oxygen consumption was presented (31,61,62). The presented configuration offers the benefit of avoiding monitoring O_2_ consumption and CO_2_ production through laboratory device, instead allowing for the application of less sophisticated respiratory monitoring techniques, such as IP. Simultaneous acquisition of both ECG and IP can be performed using e.g., Pneumonitor device, which is a recently developed device, designed for research in the fields of physiology and sports medicine (12,13,63). Thus, all the cardiorespiratory features under current study could be obtained using Pneumonitor without any additional equipment.

There are several limitations of the study. First of all, the raw ECG/RR-intervals signals and raw respiratory curves were unavailable, and thus more sophisticated parameters and parameters from information and causal domains, which could provide additional insights into the predictive models could have not been calculated. Moreover, the sample size in this study was limited, as only 369 recordings from the initial database of 992 CPET recordings were used for analysis after applying exclusion criteria based on outlier detection methods and visual inspection of the signals. Furthermore, the dataset was imbalanced in terms of patients’ sex as there were 275 men and 52 women. A larger and more balanced dataset could prove beneficial for ML model training. There was also lack of information about the amount of sport activity undertaken by the participants, which might introduce inconsistency in the study population. Additionally, one approach of age-predicted HR_max_ calculation and one threshold of HR_max_ were introduced. Some of these limitations could be overcome by the usage of the Pneumonitor device, which allows for the simultaneous acquisition of raw ECG and IP signals (63). Thanks to this, the pulmonary activity (including RespRate and VE) can be monitored without the usage of sophisticated apparatus for gas analysis and tight-fitting masks may stress some groups of patients (e.g., children). Future studies may explore the optimal percentage of HR_max_ as well as other than treadmill forms of cardiopulmonary exercise tests in order to determine the optimal settings for the prediction of VO_2peak_ for clinical practice.

This study expands the discussion on predicting cardiorespiratory fitness by highlighting the important role of submaximal testing and incorporating respiratory signals in the prediction process. The presented analysis indicates that the inclusion of respiratory parameters might improve the quality of the VO_2peak_ prediction. The use of a submaximal test based on age-predicted HR_max_ and the utilization of cardiological and respiratory parameters that can be obtained without specialized CPET equipment is an advantage of the presented approach and facilitates its potential application in clinical practice.

## Data Availability

All files are available from the Physionet database: https://physionet.org/content/treadmill-exercise-cardioresp/1.0.1/ (DOI: 10.13026/7ezk-j442)

## 5. Supporting information

Feature names presented in Figures 7 and 8 are explained in the S1 Appendix.

## 6. Author Contributions

**Conceptualization:** Maciej Rosoł, Monika Petelczyc

**Data Curation:** Maciej Rosoł, Monika Petelczyc

**Formal Analysis:** Maciej Rosoł

**Methodology:** Maciej Rosoł, Monika Petelczyc, Jakub S. Gąsior, Marcel Młyńczak

**Software:** Maciej Rosoł

**Visualization:** Maciej Rosoł

**Writing – Original Draft Preparation:** Maciej Rosoł

**Writing – Review & Editing:** Maciej Rosoł, Monika Petelczyc, Jakub S. Gąsior, Marcel Młyńczak

## 7. Acknowledgments

Research was founded by POB Biotechnology and biomedical engineering of Warsaw University of Technology within the Excellence Initiative: Research University (IDUB) programme.

## References

1. Lee J, Zhang XL. Physiological determinants of VO2max and the methods to evaluate it: A critical review. Vol. 36, Science and Sports. 2021.

2. Laukkanen JA, Rauramaa R, Salonen JT, Kurl S. The predictive value of cardiorespiratory fitness combined with coronary risk evaluation and the risk of cardiovascular and all-cause death. J Intern Med. 2007;262(2).

3. Laukkanen JA, Kurl S, Salonen R, Rauramaa R, Salonen JT. The predictive value of cardiorespiratory fitness for cardiovascular events in men with various risk profiles: A prospective population-based cohort study. Eur Heart J. 2004;25(16).

4. Jones LW, Watson D, Herndon JE, Eves ND, Haithcock BE, Loewen G, et al. Peak oxygen consumption and long-term all-cause mortality in nonsmall cell lung cancer. Cancer. 2010;116(20).

5. Bernal W, Martin-Mateos R, Lipcsey M, Tallis C, Woodsford K, McPhail MJ, et al. Aerobic capacity during cardiopulmonary exercise testing and survival with and without liver transplantation for patients with chronic liver disease. Liver Transplantation. 2014;20(1).

6. Snowden CP, Prentis JM, Anderson HL, Roberts DR, Randles D, Renton M, et al. Submaximal cardiopulmonary exercise testing predicts complications and hospital length of stay in patients undergoing major elective surgery. Ann Surg. 2010;251(3).

7. Schabort EJ, Killian SC, St Clair Gibson A, Hawley JA, Noakes TD. Prediction of triathlon race time from laboratory testing in national triathletes. Med Sci Sports Exerc. 2000;32(4).

8. Billat VL, Demarle A, Slawinski J, Paiva M, Koralsztein JP. Physical and training characteristics of top-class marathon runners. Med Sci Sports Exerc. 2001;33(12).

9. Staib JL, Im J, Caldwell Z, Rundell KW. Cross-Country Ski Racing Performance Predicted by Aerobic and Anaerobic Double Poling Power. J Strength Cond Res. 2000;14(3).

10. Sutterfield SL, Alexander AM, Hammer SM, DIdier KD, Caldwell JT, Barstow TJ, et al. Prediction of Planetary Mission Task Performance for Long-Duration Spaceflight. Med Sci Sports Exerc. 2019;51(8).

11. Levett DZH, Jack S, Swart M, Carlisle J, Wilson J, Snowden C, et al. Perioperative cardiopulmonary exercise testing (CPET): consensus clinical guidelines on indications, organization, conduct, and physiological interpretation. Br J Anaesth. 2018;120(3).

12. Mlynczak M, Zylinski M, Niewiadomski W, Cybulski G. Ambulatory Devices Measuring Cardiorespiratory Activity with Motion. In: BIODEVICES 2017 -10th International Conference on Biomedical Electronics and Devices, Proceedings; Part of 10th International Joint Conference on Biomedical Engineering Systems and Technologies, BIOSTEC 2017. 2017.

13. Młyńczak MC, Niewiadomski W, Zyliński M, Cybulski GP. Ambulatory impedance pneumography device for quantitative monitoring of volumetric parameters in respiratory and cardiac applications. In: Computing in Cardiology. 2014.

14. Petelczyc M, Kotlewski M, Bruhn S, Weippert M. Maximal oxygen uptake prediction from submaximal bicycle ergometry using a differential model. Sci Rep [Internet]. 2023;13(1):11289. Available from: 10.1038/s41598-023-38089-7

15. Marsico A, Corso SD, de Carvalho EF, Arakelian V, Phillips S, Stirbulov R, et al. A more effective alternative to the 6-minute walk test for the assessment of functional capacity in patients with pulmonary hypertension. Eur J Phys Rehabil Med. 2021;57(4).

16. Alves R, Lima MM, Fonseca C, Dos Reis R, Figueiredo PH, Costa H, et al. Peak oxygen uptake during the incremental shuttle walk test in a predominantly female population with Chagas heart disease. Eur J Phys Rehabil Med. 2016;52(1).

17. Buttar KK, Saboo N, Kacker S. A Review: Maximal Oxygen Uptake (VO2 Max) and Its Estimation Methods. International Journal of Physical Education, Sports and Health. 2019;6(6).

18. Lima LP, Leite HR, de Matos MA, Neves CDC, da Silva Lage VK, da Silva GP, et al. Cardiorespiratory fitness assessment and prediction of peak oxygen consumption by Incremental Shuttle Walking Test in healthy women. PLoS One. 2019;14(2).

19. Selland CA, Kelly J, Gums K, Meendering JR, Vukovich M. A Generalized Equation for Prediction of VO 2peak from a Step Test. Int J Sports Med. 2021;42(9).

20. Cureton KJ, Sloniger MA, O’Bannon JP, Black DM, McCormack WP. A generalized equation for prediction of VO2peak from 1-mile run/walk performance. Med Sci Sports Exerc. 1995;27(3).

21. Cooper KD, Shafer AB. Validity and Reliability of the Polar A300’s Fitness Test Feature to Predict VO2max. Int J Exerc Sci. 2019;12(4).

22. Hernandez B, Roberts B, Kodidhi A, Roelle L, Miller N, Littell LM, et al. Evaluating accuracy of estimated VO2max with wrist worn Polar Ignite compared to peak VO2 on formal cardiopulmonary exercise testing in healthy and fontan pediatric patients. J Am Coll Cardiol. 2023;81(8).

23. Chirico D, Davidson TW, Terada T, Scott K, Keast ML, Reid RD, et al. Using the 6-min walk test to monitor peak oxygen uptake response to cardiac rehabilitation in patients with heart failure. J Cardiopulm Rehabil Prev. 2020;40(6).

24. Mandic S, Walker R, Stevens E, Nye ER, Body D, Barclay L, et al. Estimating exercise capacity from walking tests in elderly individuals with stable coronary artery disease. Disabil Rehabil. 2013;35(22).

25. Wiecha S, Kasiak PS, Szwed P, Kowalski T, Cieśliński I, Postuła M, et al. VO2max prediction based on submaximal cardiorespiratory relationships and body composition in male runners and cyclists: a population study. Löllgen H, Barton M, Löllgen H, Knechtle B, editors. Elife [Internet]. 2023;12:e86291. Available from: 10.7554/eLife.86291

26. Ashfaq A, Cronin N, Müller P. Recent advances in machine learning for maximal oxygen uptake (VO2 max) prediction: A review. Vol. 28, Informatics in Medicine Unlocked. 2022.

27. Hedge ET, Amelard R, Hughson RL. Prediction of oxygen uptake kinetics during heavy-intensity cycling exercise by machine learning analysis. J Appl Physiol. 2023;134(6).

28. Szijarto A, Tokodi M, Fabian A, Lakatos BK, Shiida K, Tolvaj M, et al. Deep-learning based prediction of peak oxygen uptake in athletes using 2D echocardiographic videos. Eur Heart J Cardiovasc Imaging [Internet]. 2023 Jun 1;24(Supplement_1):jead119.244. Available from: 10.1093/ehjci/jead119.244

29. Zignoli A, Fornasiero A, Ragni M, Pellegrini B, Schena F, Biral F, et al. Estimating an individual’s oxygen uptake during cycling exercise with a recurrent neural network trained from easy-to-obtain inputs: A pilot study. PLoS One. 2020;15(3).

30. Amelard R, Hedge ET, Hughson RL. Temporal convolutional networks predict dynamic oxygen uptake response from wearable sensors across exercise intensities. NPJ Digit Med. 2021;4(1).

31. Wang Z, Zhang Q, Lan K, Yang Z, Gao X, Wu A, et al. Enhancing instantaneous oxygen uptake estimation by non-linear model using cardio-pulmonary physiological and motion signals. Front Physiol. 2022;13.

32. Mongin D, Chabert C, Courvoisier DS, García-Romero J, Alvero-Cruz JR. Heart rate recovery to assess fitness: comparison of different calculation methods in a large crosssectional study. Research in Sports Medicine. 2023;31(2).

33. Mongin D, García-Romero J, Alvero-Cruz JR. <https://physionet.org/content/treadmillexercise-cardioresp/1.0.1/>. 2021. Treadmill Maximal Exercise Tests from the Exercise Physiology and Human Performance Lab of the University of Malaga (version 1.0.1) PhysioNet.

34. Robergs RA, Dwyer D, Astorino T. Recommendations for improved data processing from expired gas analysis indirect calorimetry. Sports Medicine. 2010;40(2).

35. Noonan V, Dean E. Submaximal exercise testing: Clinical application and interpretation. Vol. 80, Physical Therapy. 2000.

36. Shushan T, Lovell R, Buchheit M, Scott TJ, Barrett S, Norris D, et al. Submaximal Fitness Test in Team Sports: A Systematic Review and Meta-Analysis of Exercise Heart Rate Measurement Properties. Vol. 9, Sports Medicine -Open. 2023.

37. Leopold E, Tuller T, Scheinowitz M. A computational predictor of the anaerobic mechanical power outputs from a clinical exercise stress test. PLoS One. 2023;18(5 MAY).

38. Chen T, Guestrin C. XGBoost: A scalable tree boosting system. In: Proceedings of the ACM SIGKDD International Conference on Knowledge Discovery and Data Mining. 2016.

39. Pedregosa F, Varoquaux G, Gramfort A, Michel V, Thirion B, Grisel O, et al. Scikitlearn: Machine learning in Python. Journal of Machine Learning Research. 2011;12.

40. Chollet F. Keras (2015). URL http://kerasio. 2017;

41. Baniecki H, Kretowicz W, Piatyszek P, Wisniewski J, Biecek P. dalex: Responsible machine learning with interactive explainability and fairness in python. Journal of Machine Learning Research. 2021;22.

42. Mart MF, Ely EW, Tolle JJ, Patel MB, Brummel NE. Physiologic responses to exercise in survivors of critical illness: an exploratory pilot study. Intensive Care Medicine Experimental. 2022;10(1).

43. Baldasseroni S, Silverii MV, Pratesi A, Burgisser C, Orso F, Lucarelli G, et al. Cardiac Rehabilitation in Advanced aGE after PCI for acute coronary syndromes: predictors of exercise capacity improvement in the CR-AGE ACS study. Aging Clin Exp Res. 2022;

44. Izquierdo MC, Lopes S, Teixeira M, Polónia J, Alves AJ, Mesquita-Bastos J, et al. The Chester step test is a valid tool to assess cardiorespiratory fitness in adults with hypertension: reducing the gap between clinical practice and fitness assessments. Vol. 42, Hypertension Research. 2019.

45. Garcia-Tabar I, Iturricastillo A, Castellano J, Cadore EL, Izquierdo M, Setuain I. Predicting Cardiorespiratory Fitness in Female Soccer Players: The Basque Female Football Cohort Study. Int J Sports Physiol Perform. 2022;17(1).

46. Altmann S, Neumann R, Härtel S, Woll A, Buchheit M. Using submaximal exercise heart rate for monitoring cardiorespiratory fitness changes in professional soccer players: A replication study. Int J Sports Physiol Perform. 2021;16(8).

47. Sartor F, Vernillo G, De Morree HM, Bonomi AG, La Torre A, Kubis HP, et al. Estimation of maximal oxygen uptake via submaximal exercise testing in sports, clinical, and home settings. Vol. 43, Sports Medicine. 2013.

48. Albouaini K, Egred M, Alahmar A, Wright DJ. Cardiopulmonary exercise testing and its application. Vol. 83, Postgraduate Medical Journal. 2007.

49. Klaren RE, Horn GP, Fernhall B, Motl RW. Accuracy of the VO2peak prediction equation in firefighters. Journal of Occupational Medicine and Toxicology. 2014;9(1).

50. Mahar MT, Welk GJ, Rowe DA, Crotts DJ, McIver KL. Development and Validation of a Regression Model to Estimate VO2peak from PACER 20-m Shuttle Run Performance. J Phys Act Health. 2016;3(2).

51. Loe H, Nes BM, Wisløff U. Predicting VO2peak from submaximal-and peak exercise models: The HUNT 3 fitness study, Norway. PLoS One. 2016;11(1).

52. Harmsen WJ, Ribbers GM, Slaman J, Heijenbrok-Kal MH, Khajeh L, van Kooten F, et al. The six-minute walk test predicts cardiorespiratory fitness in individuals with aneurysmal subarachnoid hemorrhage. Top Stroke Rehabil. 2017;24(4).

53. Billinger SA, Van Swearingen E, Mcclain M, Lentz AA, Good MB. Recumbent Stepper Submaximal Exercise Test to Predict Peak Oxygen Uptake. Med Sci Sports Exerc [Internet]. 2012;44(8). Available from: <https://journals.lww.com/acsm-msse/Fulltext/2012/08000/Recumbent_Stepper_Submaximal_Exercise_Test_to.17.asp>;x

54. Herda AA, Lentz AA, Mattlage AE, Sisante JF, Billinger SA. Cross-validation of the recumbent stepper submaximal exercise test to predict peak oxygen uptake in older adults. Phys Ther. 2014;94(5).

55. Deka P, Pozehl BJ, Pathak D, Williams M, Norman JF, Alonso WW, et al. Predicting maximal oxygen uptake from the 6 min walk test in patients with heart failure. ESC Heart Fail. 2021;8(1).

56. Mlynczak M. Temporal orders and causal vector for physiological data analysis. In: Proceedings of the Annual International Conference of the IEEE Engineering in Medicine and Biology Society, EMBS. 2020.

57. Rosoł M, Młyńczak M, Cybulski G. Granger causality test with nonlinear neuralnetwork-based methods: Python package and simulation study. Comput Methods Programs Biomed. 2022;216.

58. Schulz S, Adochiei FC, Edu IR, Schroeder R, Costin H, Bär KJ, et al. Cardiovascular and cardiorespiratory coupling analyses: A review. Vol. 371, Philosophical Transactions of the Royal Society A: Mathematical, Physical and Engineering Sciences. 2013.

59. Rosol M, Gasior JS, Walecka I, Werner B, Cybulski G, Mlynczak M. Causality in cardiorespiratory signals in pediatric cardiac patients. Annu Int Conf IEEE Eng Med Biol Soc. 2022;2022.

60. Wipf D, Nagarajan S. A new view of automatic relevance determination. In: Advances in Neural Information Processing Systems 20 -Proceedings of the 2007 Conference. 2008.

61. Gastinger S, Sorel A, Nicolas G, Gratas-Delamarche A, Prioux J. A comparison between ventilation and heart rate as indicator of oxygen uptake during different intensities of exercise. J Sports Sci Med. 2010;9(1).

62. Beltrame T, Amelard R, Wong A, Hughson RL. Prediction of oxygen uptake dynamics by machine learning analysis of wearable sensors during activities of daily living. Sci Rep. 2017;7.

63. Gąsior JS, Młyńczak M, Rosoł M, Wieniawski P, Walecka I, Cybulski G, et al. Validity of the Pneumonitor for RR intervals acquisition for short-term heart rate variability analysis extended with respiratory data in pediatric cardiac patients. Kardiol Pol. 2023;81(5).

